# Bayesian network enables interpretable and state-of-the-art prediction of immunotherapy responses in cancer patients

**DOI:** 10.1101/2022.11.02.22281835

**Authors:** Hideki Hozumi, Hideyuki Shimizu

## Abstract

Immune checkpoint inhibitors, especially PD-1/PD-L1 blockade, have revolutionized cancer treatment and brought tremendous benefits to patients who otherwise would have had a limited prognosis. Nonetheless, only a small fraction of patients responds to immunotherapy, and the costs and side effects of immune checkpoint inhibitors cannot be ignored. With the advent of machine and deep learning, clinical and genetic data has been used to stratify patient responses to immunotherapy. Unfortunately, these approaches have typically been “black-box” methods that are unable to explain their predictions, thereby hindering their clinical and responsible application. Herein, we developed a “white-box” Bayesian network model that achieves accurate and interpretable predictions of immunotherapy responses against non-small cell lung cancer (NSCLC). This Tree-Augmented naïve Bayes model (TAN) precisely predicted durable clinical benefits and distinguished two clinically significant subgroups with distinct prognoses. Furthermore, Our state-of-the-art white-box TAN approach achieved greater accuracy than previous methods. We hope our model will guide clinicians in selecting NSCLC patients who truly require immunotherapy and expect our approach will be easily applied to other types of cancer.

**Structured Abstract:** *Background:* Immune checkpoint inhibitors have revolutionized cancer treatment. Given that only a small fraction of patients responds to immunotherapy, patient stratification is a pressing concern. Unfortunately, the “black-box” nature of most of the proposed stratification methods, and their far from satisfactory accuracy, has hindered their clinical application.

*Method:* We developed a “white-box” Bayesian network model, with interpretable architecture, that can accurately predict immunotherapy response against non-small cell lung cancer (NSCLC). We collected clinical and genetic information from several independent studies, and integrated this via the Tree-Augmented naïve Bayes (TAN) approach.

*Findings:* This TAN model precisely predicted durable clinical benefit and distinguished two clinically significant subgroups with distinct prognoses, achieving state-of-the-art performance than previous methods. We also verified that TAN succeeded in detecting meaningful interactions between variables from data-driven approach. Moreover, even when data have missing values, TAN successfully predicted their prognosis.

*Interpretation:* Our model will guide clinicians in selecting NSCLC patients who genuinely require immunotherapy. We expect this approach to be easily applied to other types of cancer. To accelerate the uptake of personalized medicine via access to accurate and interpretable models, we provide a web application (https://pred-nsclc-ici-bayesian.shinyapps.io/Bayesian-NSCLC/) for use by the researchers and clinicians community.

*Funding:* KAKENHI grant from the Japan Society for the Promotion of Science (JSPS) to H.S (21K17856).

## Introduction

Lung cancer is the most prevalent cancer and the leading cause of cancer-related death in men and women worldwide^1^. Non-small cell lung cancer (NSCLC) accounts for nearly 85 percent of all lung cancers, and its five-year survival rate remains dismal, ranging from 68% in patients with stage IB cancer to as low as 10% in patients with stage IVA-IVB cancer^2^. Since the invention of immune checkpoint inhibitors (ICIs), many patients have gained tremendous benefits, with improved life expectancy^3^. For instance, nivolumab, an inhibitor of the programmed cell death 1 (PD-1)/ligand (PD-L1) pathway, increased the 2-year survival rate of patients with stage IIIB/IV cancer from 16% to approximately 30%^4^.

The decision to administer ICIs to NSCLC patients has been based primarily on the expression level of PD-L1 on the surface of cancer cells, referred to as PD-L1 score^5^. In most cases, patients with higher PD-L1 scores are deemed suitable candidates for ICIs. Nonetheless, numerous studies have demonstrated that not all patients with higher PD-L1 scores respond to ICIs, and even patients with lower PD-L1 scores respond to ICIs^5–7^. According to a meta-analysis of randomized controlled trials, PD-L1 expression alone was insufficient to predict immunotherapy response^7^. In support of this, the PD-L1-based predictive ability was reported to be 0.646 (based on the area under the curve, AUC)^8^, indicating that other factors must determine immunotherapy benefits. Further, immunotherapy can have devastating side-effects, particularly immune-related adverse events such as pancreatitis and interstitial pneumonia^9^. The use of ICIs in patients who do not respond to treatment may thus eventually reduce their life expectancy. It is therefore urgent to elucidate the factors other than PD-L1 score that determine the response and prognosis of patients under immunotherapy^10^.

Studies to identify factors for stratifying NSCLC patients on ICI treatment have focused on the tumor mutational burden (TMB). Tumors with high TMB would contain many neoantigens and generally respond well to ICIs^10^. However, the predictive ability of TMB was 0.601, based on AUC^8^. Rather than relying on a single indicator (such as PD-L1 score or TMB) to predict immunotherapy response, methods combining multiple factors have emerged. For example, LIPI^11^ and EPSILoN^12^ integrate clinical data such as clinical stage, performance status, and smoking status. The ratio of neutrophils to lymphocytes, a predictor of rapid progression^10^, has been incorporated into these methods. Despite the use of multiple variables, prediction of immunotherapy response rate has remained inadequate, with AUC values of 0.606 and 0.666 for LIPI and EPSILoN, respectively^13^. This evidence indicates that classical approaches cannot provide satisfactory predictions concerning immunotherapy.

In recent years, machine learning (ML)-based methods have been applied to unravel the factors determining the efficacy of ICI treatment for NSCLC. For one example, the AUC of a neural network model that integrated several factors (TMB, PD-L1 score, mutant-allele tumor heterogeneity, and immune-related pathways) was as high as 0.80 in a test cohort^14^. Another study integrated PD-L1 score and CT images achieved an AUC of 0.76^15^. Other ML methods, such as LightGBM, XGBoost, and regression analysis have also been investigated^16^. Although they harness various types of information (PD-L1 score, radiological images, and clinical features) as input, the AUC remains below 0.80 even for their best models, indicating that it remains challenging to predict responses in ICI therapy. Moreover, ML methods, including neural networks, often lack transparency due to the complexity arising from neural connections and mathematical abstractions^17–19^, making it potentially impossible to explain their predictions. This “black-box” nature has retarded the clinical application of established models. Predictive models with higher accuracy and accountability are therefore necessary for the appropriate use of ICIs in NSCLC patients.

With this in mind, we harnessed Bayesian theory and developed an interpretable AI model with state-of-the-art predictive power about immunotherapy. Specifically, we utilized Bayesian network (BN)-based models that capitulate causal relationships in the form of a graphical model^20^, allowing us to avoid the black-box problems prevalent in other ML methods^17^. We demonstrate that a Tree-Augmented naïve Bayes (TAN) model predicts the durable clinical benefit (DCB) of patients treated with ICIs with comparable or even better accuracy than conventional ML methods, stratifying patients in a clinically significant manner. It achieved robust predictive ability, even with limited information. This data-driven approach can be used to further elucidate the factors determining immunotherapeutic responses. We anticipate that our interpretable and state-of-the-art approach will expand the knowledge of immunotherapy and be readily applicable to other types of cancer.

## Methods

### Public cohorts

The cBioPortal (http://www.cbioportal.org)^21^ was accessed to retrieve clinical and mutation data for NSCLC patients. We chose two studies examining the effects of ICIs on NSCLC patients^8,22^ to use as a dataset for this study. The inclusion criteria and clinical and mutation information for the two cohorts are explained in the original papers^8,22^.

The characteristics of our dataset are shown in Table 1, including age (<65 years or not), gender, smoking status, and histopathological information. We excluded 25 samples, comprising mostly those with unspecified histological data (described only as “NSCLC”), and a few categorized as “Large Cell Neuroendocrine Carcinoma”. We obtained mutation data to prepare the “frequency-based” and “evidence-based” gene-sets. For the patients in these cohorts, we also analyzed progression-free status.

**Table 1.**
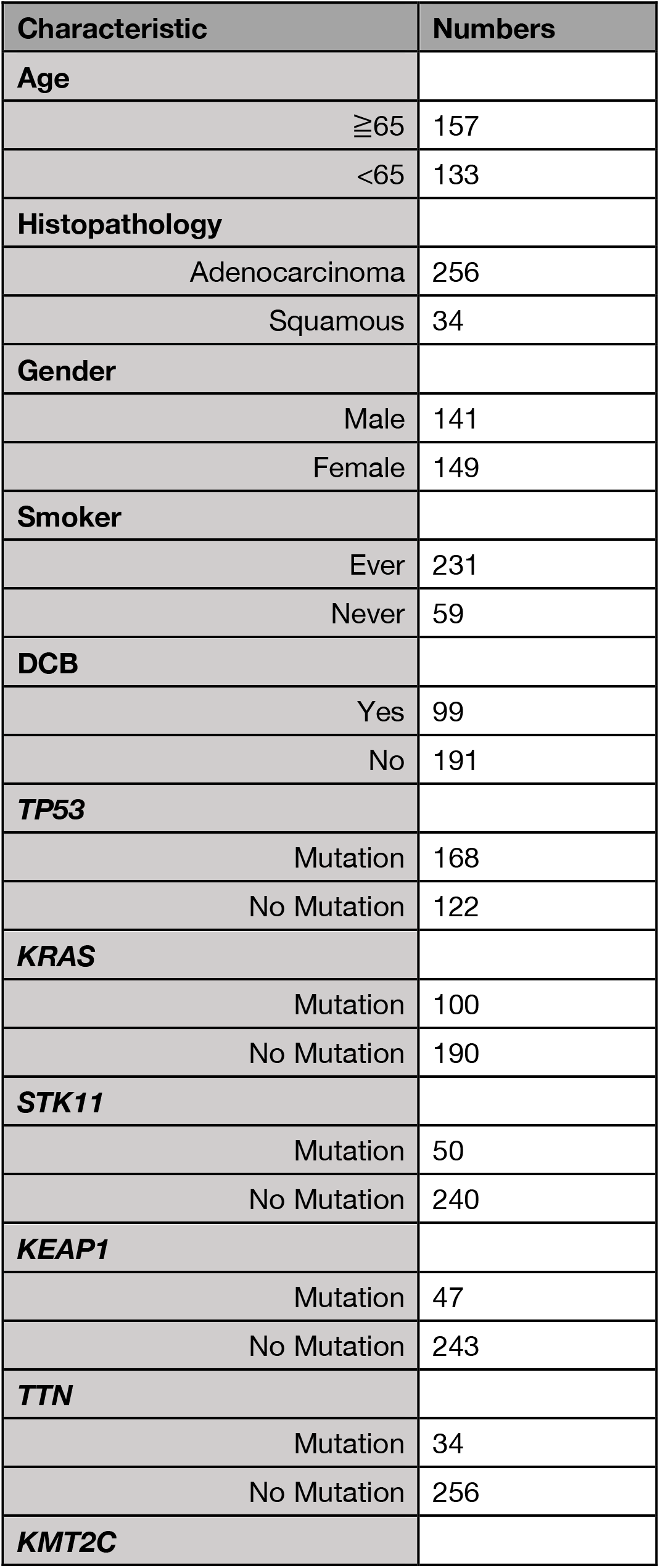
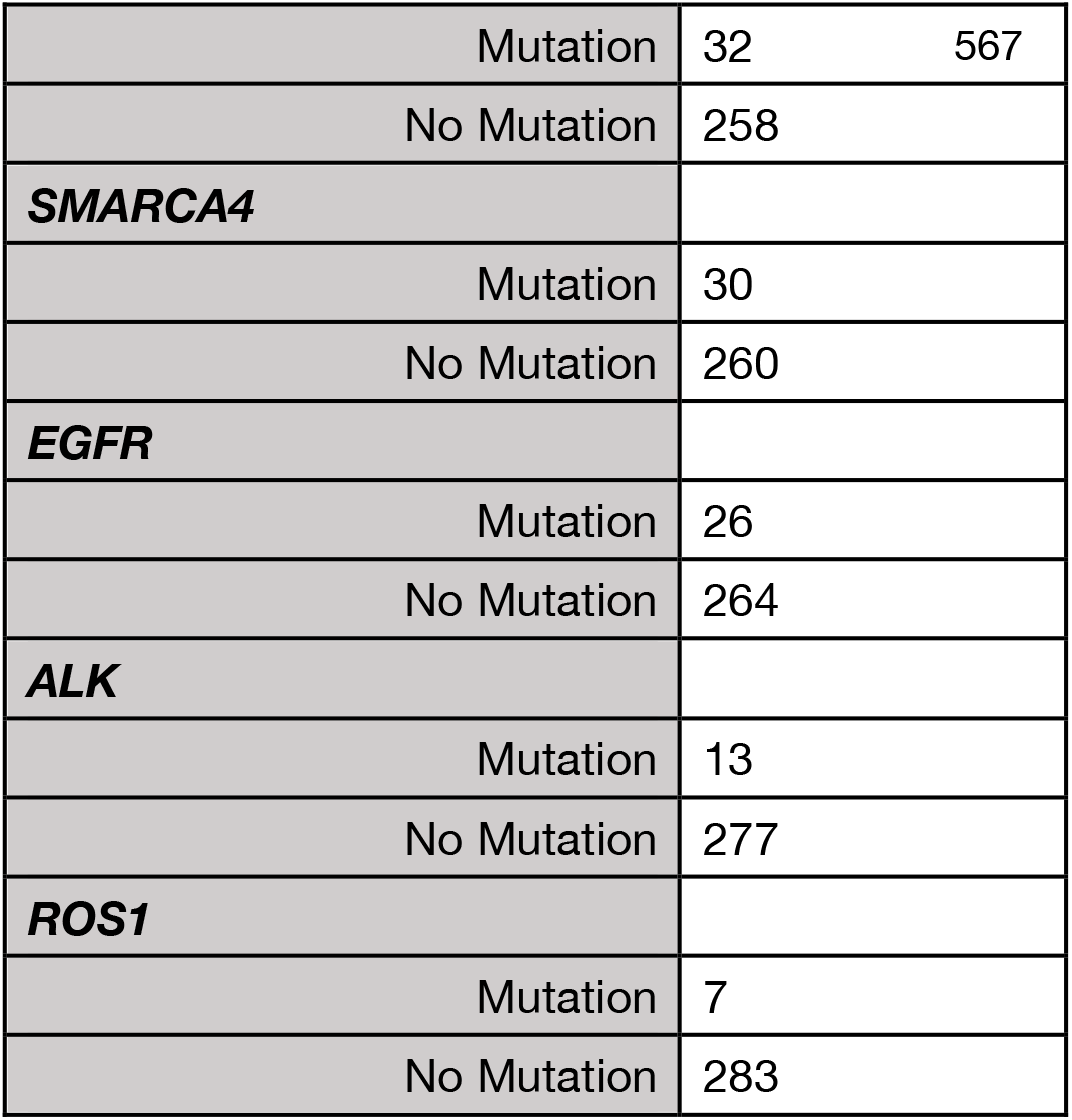
Characteristics of the dataset.

### Model construction

The characteristic of TAN lies in its structural constraints that each explanatory variable can be connected with one node other than an objective variable.

A complete undirected graph with nodes and edges is constructed to estimate this structure. In this stage, one node is wholly connected to every other node. Each variable is described as *X*_1_, …, *X*_*n*_, and mutual information values are given to each edge. The edge weights between two nodes (*X*_*i*_, *X*_*j*_) are given by Equation 1:

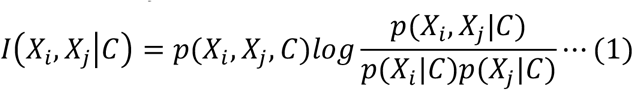

To obtain the constrained structure of TAN from this complete graph, a structure with the highest total weights under the constraint is used as an estimated structure. To transform the given undirected graph tree into a directed one, a root variable is randomly chosen, and the direction of the edges is set to outward from the root variable^20,23,24^. The data were then randomly split into training (2/3) and test data (1/3) (Figure 1b).

**Figure 1.**
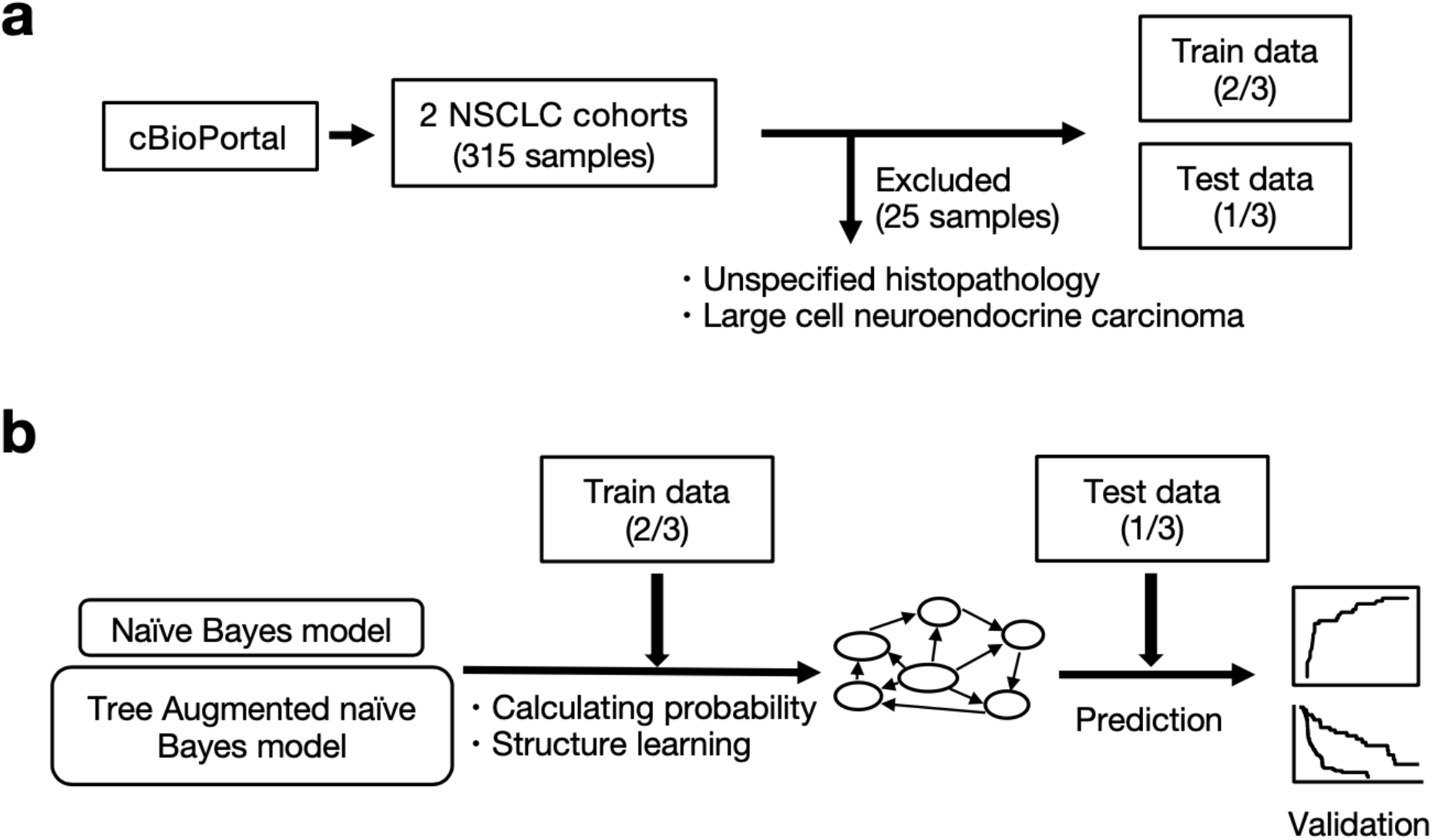
Workflow of the study. (**a**) We obtained clinical and genetic data of non-small cell lung cancer (NSCLC) patients from cBioPortal (http://www.cbioportal.org). There were 315 data samples, of which 25 samples were excluded because they had insufficient histopathology data, or because the disease were rare. Two-thirds of the data was used to construct the models (train data), and the rest was used for evaluation (test data). (**b**) We developed the naïve Bayes (NB) and Tree-Augmented naïve Bayes (TAN) models and evaluated their predictive accuracy for whether patients will benefit from immunotherapy. We performed survival analyses to compare the two groups based on the binary predictions by TAN model.

The training data were used to construct the models and to learn the conditional probability between each node. ROC curves were constructed from the test-data predictions. We constructed the model using the bnlearn (4.7.1) R package, and used the ROCR package (1.0-11) for evaluation.

### Model evaluation

Model-averaging methods were adopted to measure the reliability of the connections between nodes in the network, by performing multiple structural estimations using the hill-climbing method^23^. In the Bayesian network, it is important to measure the confidence level for a particular graph feature (the graph edge). This confidence level (in terms of relative frequencies) referred to as arc strength^23,25,26^, is defined as the number of times an internode connection appears while generating multiple graphs; frequencies >85% are considered strong^23^.

We adopted two model-averaging methods for evaluating the node connections of our model. The first is the bootstrap approach, which applies nonparametric bootstrapping to generate multiple networks, and estimates the arc strength^23,25^.

Algorithm 1 provides the specific method.

#### Algorithm 1

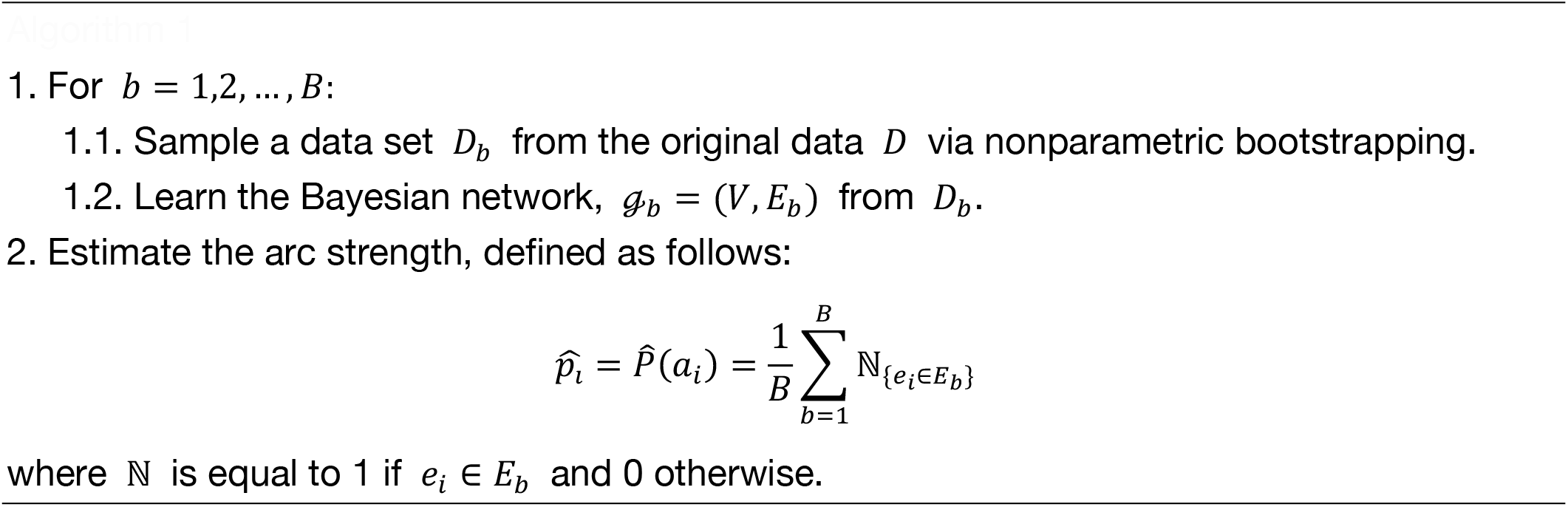

The second model-averaging method is the random generation of multiple graphs from a uniform distribution, using the MCMC algorithm (Algorithm 2). We randomly sampled one graph for every 50 graphs generated and measured arc strength from 500 sampled graphs^26^.

#### Algorithm 2

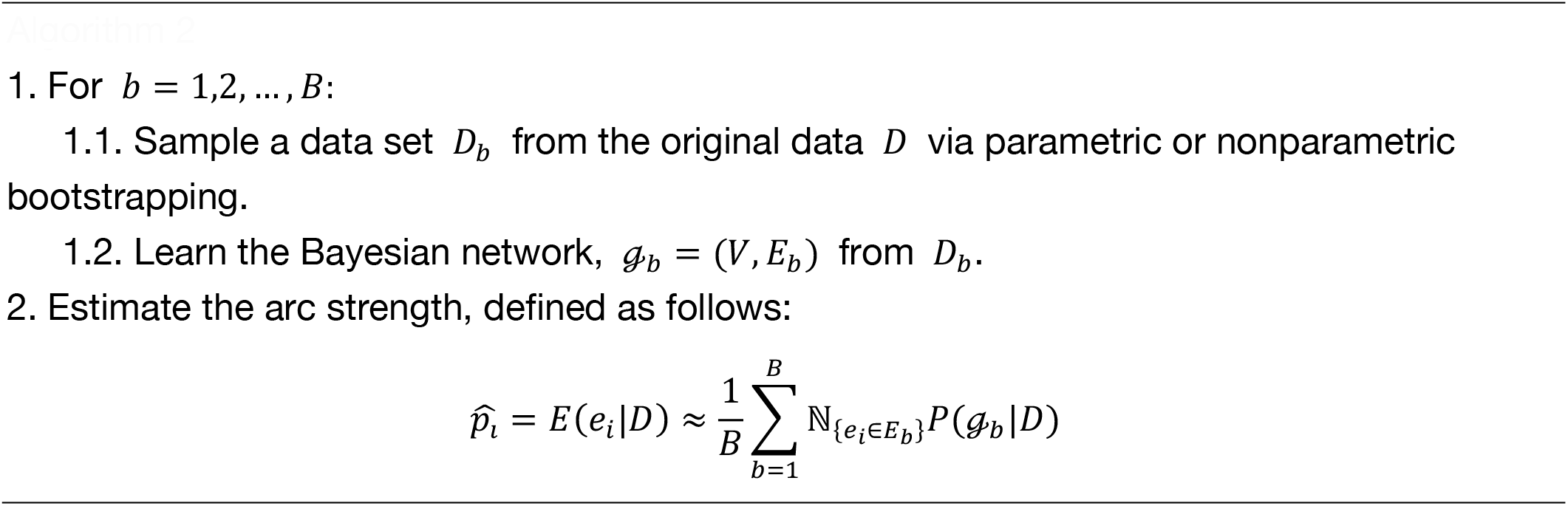

The robustness of the TAN structure estimation was evaluated by examining whether the connections between nodes determined to be significant by these model-averaging methods were also present in the structure of the TAN.

### Inference with limited evidence

To estimate the conditional probability of an event using only the limited evidence available, we used the cpquery function of the bnlearn package (4.7.1). In this method, logic sampling, or an approximate inference, enables it to obtain the probability^27^. First, a new data set is created by randomly extracting data that matches the specified evidence from the whole data set. In our case, patient profiles or genetic mutation information were specified. By repeating this method, one million random samples are generated, and an approximate probability is returned based on them.

### Survival analysis

We conducted survival analysis using the survival package (3.3-1). *P*< 0.05 was considered statistically significant.

### Data availability

All clinical and mutation information is available from the cBioPortal database, (http://www.cbioportal.org), and the specific explanation of each cohort can be obtained in the original papers^8,22^.

We provide a web application (https://pred-nsclc-ici-bayesian.shinyapps.io/Bayesian-NSCLC/) using the shiny package (1.7.2), providing both frequency- and evidence-based models.

### Code availability

The R code for training the NB and TAN models, and for validation and scoring via ROC and survival analysis are available at GitHub (https://github.com/Hideki-Hozumi/Prediction-with-bayesian-network.).

## Results

### Manual curation of clinical information related to immunotherapy

To develop a state-of-the-art explanatory model, we first retrieved data for immunotherapy-receiving cancer patients from cBioPortal (http://www.cbioportal.org), which offers clinical data with mutational information^21^. Specifically, two previously published studies^8,22^ examining the effect of ICIs on NSCLC patients were selected and used as a dataset for this study: the Hellman cohort, comprising of 75 NSCLC patients who underwent immunotherapy^22^, and the Rivzi cohort, of 240 NSCLC patients treated with immunotherapy^8^. In total, our dataset includes 315 patients (Figure 1a). The characteristics of our dataset are shown in Table 1.

Among the available clinical information, we set the objective variable as DCB, which is defined in the revised RECIST guideline (version 1.1) as partial response/stable disease lasting >6 months^28^. We focused on DCB because the follow-up criteria for overall survival and progression-free status were inconsistent between the two cohorts^8,22^. Given that DCB has been used to assess the efficacy of immunotherapy for various tumors such as melanoma^29^ and lung cancer^8^, we believe that predicting DCB is of clinical value for stratifying the patients in our study.

We used the three known clinical risk factors of NSCLC: age (< 65years of age or not^30^), gender, and smoking status^31^. Our model incorporated histopathological information, because the pathological subtype substantially affects the prognosis^32^. We excluded 25 samples for which there was insufficient histopathological information (Figure 1a).

Genetic covariates were determined in two ways. First, genes with mutation rates higher than 10% in our dataset were incorporated (hereafter, the “frequency-based geneset”); these include *TP53, KRAS, TTN, KMT2C, SMARCA4, STK11*, and *KEAP1*. Second, based on a literature survey, we identified six genes (*KRAS*^33^ *STK11*^34,35^, *TP53*^36^, *EGFR*^37^, *ALK*^37^, and *ROS1*^37^) associated with NSCLC-patient ICI-treatment responses or prognosis (hereafter, “evidence-based geneset”). We categorized “Deletion", “in-frame deletion (IF del)”, “frameshift deletion (FS del)”, “splice mutation (Splice)” and “Missense” modifications as “Mutation” since they would likely impair the original function of the gene^38,39^.

We attempted to decipher the relationships underlying DCB by combining clinical characteristics with mutation data. For this purpose, we randomly divided the dataset into training and test data (Figure 1a), using the former to build a model and the latter solely for evaluation^40^. Receiver operating characteristic (ROC) analysis was performed to evaluate model performance. Survival analysis was conducted to verify the model’s ability to predict prognosis in addition to DCB (Figure 1b). We describe the model construction procedure in the following section.

### Tree-Augmented naïve Bayes model robustly and interpretability predicted DCB

We harnessed a Bayesian network graphical model to achieve accurate and interpretable predictions of DCB. Bayesian networks graphically represent multivariate probability distributions^20^, and are broadly applied in various biomedical tasks, including gene network feature selection^41^, signaling network prediction^42^, and predicting hematological malignancy type^43^. Naïve Bayes (NB) networks, the simplest type of Bayesian network, but generally achieve favorable prediction accuracy^44^. Based on Bayes’ theorem (Equation 2), NB assumes that all covariates are equally important without distinction and are conditionally independent given a class value (Equation 3)^44^.

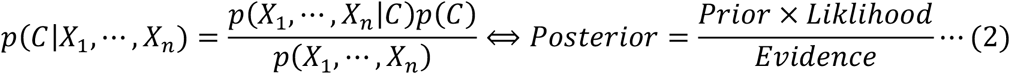

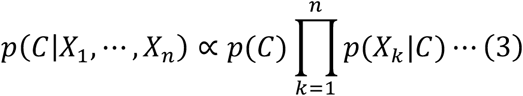

The probability associated with a parent node (objective variable) is described as *p*(*C*), and the probability is updated to *p*(*C*|*X*_1_, …, *X*_*n*_) when explanatory information from child nodes (*X*_1_, …, *X*_*n*_) is provided. In terms of its network structure, arrows (directed edges) extend from one node (a parent node or objective variable) to all other nodes (child nodes) (Supplementary Figure 1a). Despite its simple design and assumptions, NB achieves much better classification than expected, and is used in medical data analysis^45^. Nonetheless, in its original form, it depends relatively heavily on the assumption that the covariates are statistically independent, hampering its application to real-world biomedical data. To address this, we utilized Tree-Augmented naïve Bayes models (hereafter, “TAN”; Equation 4):

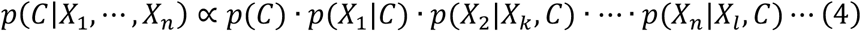

TAN alleviates the conditional independence between features, while keeping the directed acyclic graph simpler than in conventional neural network models (Supplementary Figure 1b, 1c). TAN does not assume conditional independence, partially allowing dependent relationships between variables (Equation 3)^46^. Therefore, because it can express a greater number of states, TAN must outperform NB model. Indeed, it has been applied in numerous biomedical tasks, including risk stratification in pulmonary hypertension^47^, and mammography^48^, achieving high accuracy. Here, we used NB and TAN to establish predictive models with higher accuracy and interpretability, and compared their ability to predict DCB in NSCLC patients.

First, we constructed frequency-based models, using clinical data and seven genes from the frequency-based geneset (*TP53, KRAS, TTN, KMT2C, SMARCA4, STK11*, and *KEAP1*) as covariates. The structure of frequency-based NB is shown in Figure 2a. TAN structure was estimated using a training dataset (Figure 2b). For the NB model, the area under the curve (AUC) was 0.686 for the training dataset and 0.726 for the test dataset. (Figure 2c); for the TAN model, these values were 0.836 and 0.728, respectively (Figure 2d). These results indicate that the TAN model has comparable or greater predictive accuracy than the NB model.

**Figure 2.**
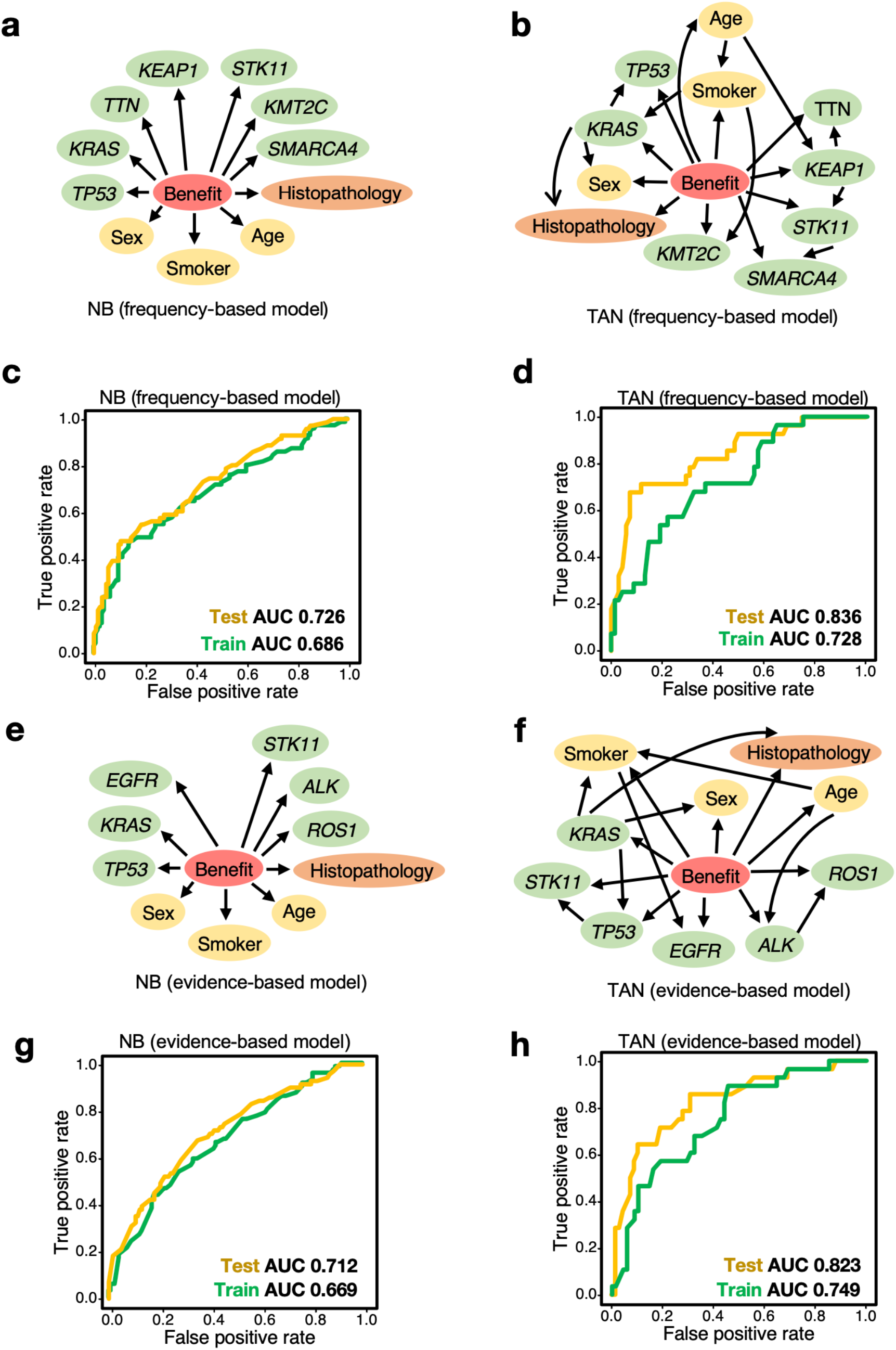
Bayesian network predicted the benefit of immunotherapy with high accuracy. (**a, b**) The naïve Bayes (NB, **a**) and Tree-Augmented naïve Bayes (TAN, **b**) models were trained using the frequency-based dataset. The predictor variable “Benefit” (DCB, shown in red) is defined in the RECIST guideline (version 1.1)^28^. Explanatory variables include patient data (yellow), tumor tissue information (orange), and the frequency-gene dataset (green). (**c, d**) Predictive performance of the frequency-based models (**a, b**) for the test dataset. TAN achieved greater accuracy than NB in terms of the Area Under the Curve (AUC), and was comparable to, or even more accurate than, state-of-the art methods^15,49,50^. (**e, f**) The naïve Bayes (NB, **e**) and Tree-Augmented naïve Bayes (TAN, **f**) models were trained using the evidence-based dataset. (**g, h**) Predictive performance of the evidence-based models (**e, f**) in the test dataset. TAN was more accurate than NB in terms of AUC, and comparable to, or even more accurate than, state-of-the-art methods^15,49,50^.

We next constructed evidence-based NB (Figure 2e) and TAN (Figure 2f) models, using clinical information and six genes from the evidence-based geneset (*KRAS, STK11, TP53, EGFR, ALK*, and *ROS1*) as covariates, using the same approach used for the frequency-based models. Using the test dataset, the NB and TAN AUCs were 0.712 (Figure 2g) and 0.823 (Figure 2h), respectively, suggesting that TAN outperformed NB.

These lines of evidence demonstrate that the optimized TAN model outperforms NB, and robustly predicts DCB via frequency- and evidence-based approaches. Its performance is comparable to that of other cutting-edge methods^15,49,50^, while retaining explainability.

### Optimized TAN yields a robust graphical structure

We next evaluated the robustness of the structural estimation of our model in immunotherapy. We statistically generated multiple directed acyclic graphs (DAGs): significant edges (internode connections) were detected when it appeared in >85% of the graphs.

We used two model-averaging methods^25,51^ to determine if the relationships identified by our methodology (Figure 2b for the frequency-based model and Figure 2f for the evidence-based model, respectively) were sufficiently robust. We performed bootstrap sampling^25^ and Markov chain Monte Carlo (MCMC) methods to randomly constructed DAGs from a uniform distribution, as previously reported^51^. This revealed several significant connections (Table 2, Figure 3a, 3b for the frequency-based model; Table 3, Figure 3c, 3d for the evidence-based model). These results demonstrate that the model-averaging methods produce similar architectures, indicating that our method robustly discovers crucial relationships governing the immunotherapy response.

**Table 2.** Node verification via bootstrapping and MCMC for the frequency-based model.

| Methods | Connection | Strength | TAN model |
| --- | --- | --- | --- |
| Bootstrap | Smoker-KRAS | 0.956 | ○ |
|  | Smoker-KMT2C | 0.874 | ○ |
|  | TP53-KRAS | 0.990 | ○ |
|  | KRAS-Cancer | 0.968 | ○ |
|  | TTN-Cancer | 0.854 | × |
|  | STK11-KEAP1 | 1.000 | ○ |
| MCMC | Cancer-KRAS | 1.000 | ○ |
|  | Smoker-Age | 0.878 | ○ |
|  | Smoker-TP53 | 0.982 | × |
|  | Smoker-KRAS | 0.982 | ○ |
|  | Smoker-KEAP1 | 0.962 | ○ |
|  | KRAS-TP53 | 1.000 | ○ |
|  | TTN-Cancer | 0.978 | × |
|  | TTN-KMT2C | 0.868 | × |
|  | TTN-SMARCA4 | 0.974 | × |
|  | STK11-TP53 | 0.978 | × |
|  | KEAP1-SMARCA4 | 0.970 | × |
|  | KEAP1-STK11 | 1.000 | ○ |

**Table 3.** Node verification via bootstrapping and MCMC for the evidence-based model.

| Methods | Connection | Strength | TAN model |
| --- | --- | --- | --- |
| **Bootstrap** | Cancer-*KRAS* | 0.978 | ○ |
|  | Cancer-*ROS1* | 0.880 | × |
|  | Smoker-*TP53* | 0.942 | × |
|  | Smoker-*EGFR* | 0.872 | ○ |
|  | *KRAS-TP53* | 1.000 | ○ |
|  | *STK11-TP53* | 0.847 | ○ |
|  | *ALK-ROS1* | 0.956 | ○ |
| **MCMC** | Smoker-*KRAS* | 0.958 | ○ |
|  | Smoker-*STK11* | 0.980 | × |
|  | Smoker-*TP53* | 0.998 | × |
|  | Smoker-*EGFR* | 0.908 | ○ |
|  | *KRAS-Cancer* | 1.000 | ○ |
|  | *KRAS-TP53* | 1.000 | ○ |
|  | *STK11-TP53* | 1.000 | ○ |
|  | *ALK-EGFR* | 0.942 | × |
|  | *ROS1-Cancer* | 1.000 | × |
|  | *ROS1-ALK* | 0.970 | ○ |

**Figure 3.**
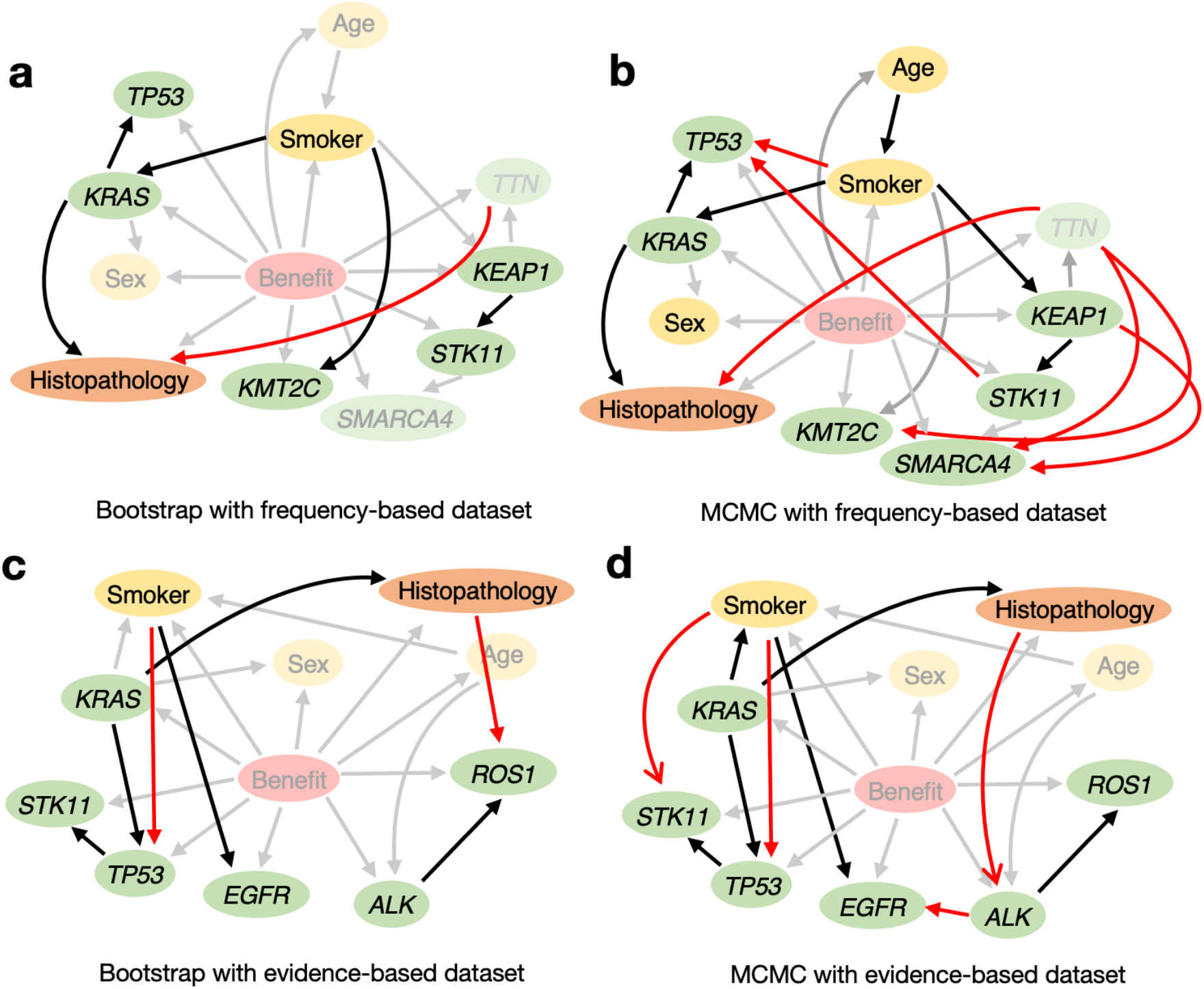
Evaluating the validity of the structure estimated by TAN through model-averaging methods. The linkages between nodes estimated by TAN were validated using model-averaging methods (bootstrap and MCMC). (**a, b**) Bootstrap (**a**) and MCMC (**b**) sampling was used to create models using frequency-based datasets; connections considered to be significant in each process are illustrated. Relationships detected by the model-averaging methods but not by TAN are shown in red. Connections detected both by model-averaging methods and TAN are in black. (**c, d**) Bootstrap (**c**) and MCMC (**d**) sampling was used to create models using evidence-based datasets; connections considered to be significant in each process are illustrated. The dependencies among variables estimated by TAN included many of the connections detected by model-averaging methods, indicating the robustness of our models. See also Tables 2 and 3.

### Our TAN model stratifies and inferences even with limited clinical information

Patient stratification is crucial to the development of personalized medicine^52^. We thus evaluated our model’s applicability to the stratification of NSCLC patients. We obtained the progression-free status of the patients in our dataset from the cBioPortal database^8,22^. Our models identified two distinct and clinically significant groups based on binary prediction (Figure 4a for the frequency-based model and Figure 4b for the evidence-based model).

**Figure 4.**
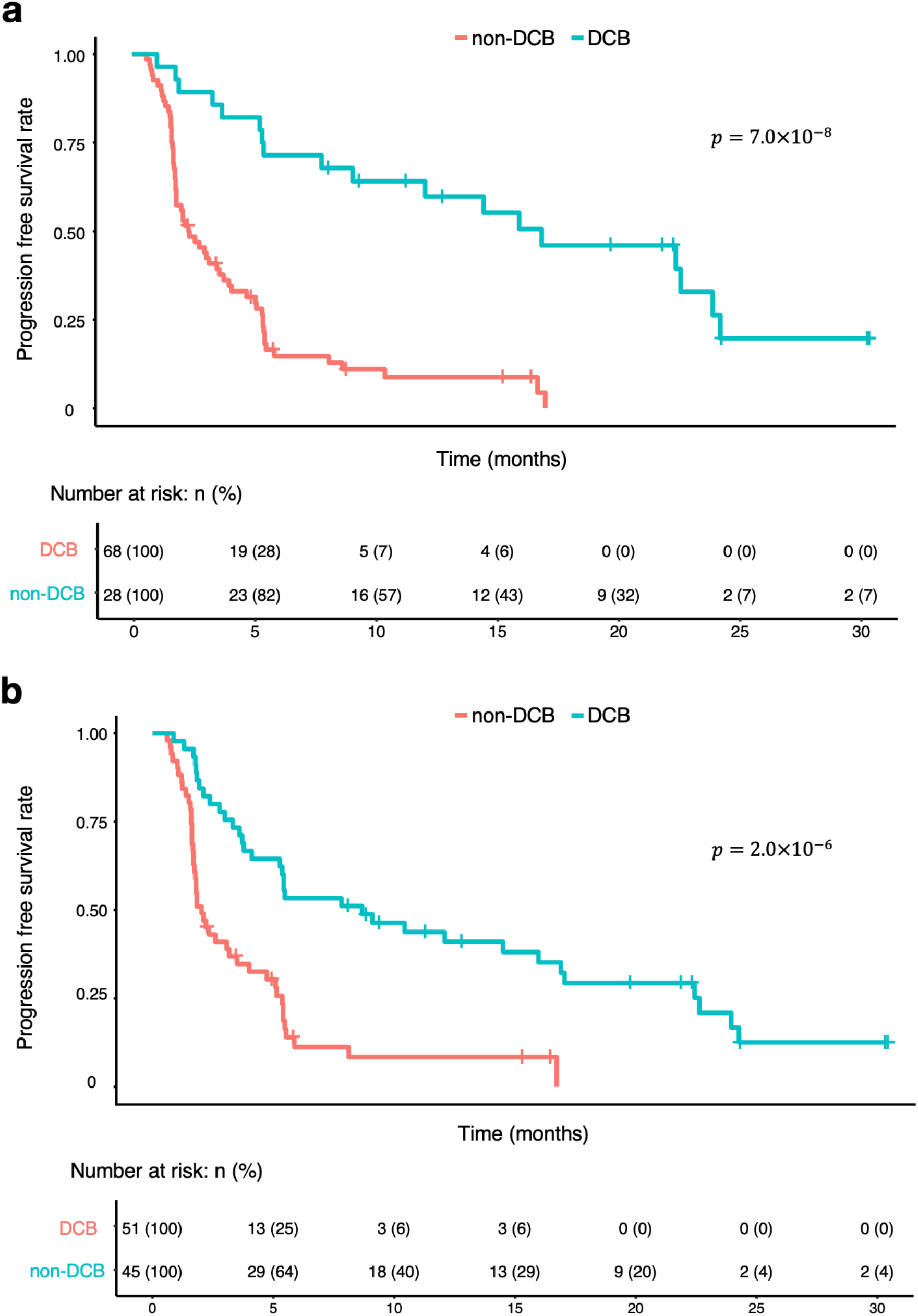
Our TAN-based interpretable models stratify NSCLC patient prognosis. (**a, b**) We tested whether these TAN models are suitable for stratifying progression-free survival. We classified patients into two groups (durable clinical benefit “DCB”, and “non-DCB”) based on the binary predictions of the frequency-based (**a**) or evidence-based (**b**) models, and estimated progression-free survival status of the patients in our dataset via the Kaplan-Meier method. The *p*-values shown in this figure are from log-rank tests.

Importantly, the optimized TAN model can handle missing data and calculate conditional probabilities. For instance, it can predict whether a tumor will respond to immunotherapy, even if all we know about a particular NSCLC patient is that they have *TP53* and *STK11* mutations; the estimated response probability is 0.163, indicating that this patient would not benefit substantially from ICIs (Figure 5). This speculation is consistent with established evidence^33^. In contrast, for a young female patient with a *KMT2C* mutation, but no *STK11* and *TP53* mutations, the estimated ICI response probability is 0.592, indicating that ICI treatment would be valuable. Previous models, including those based on ML and deep learning methods, cannot adequately handle missing data, requiring all of the necessary information^53^. Given that data acquisition can be laborious, particularly in clinical settings, our model may help clinicians in decision-making, especially in data-limited situation.

**Figure 5.**
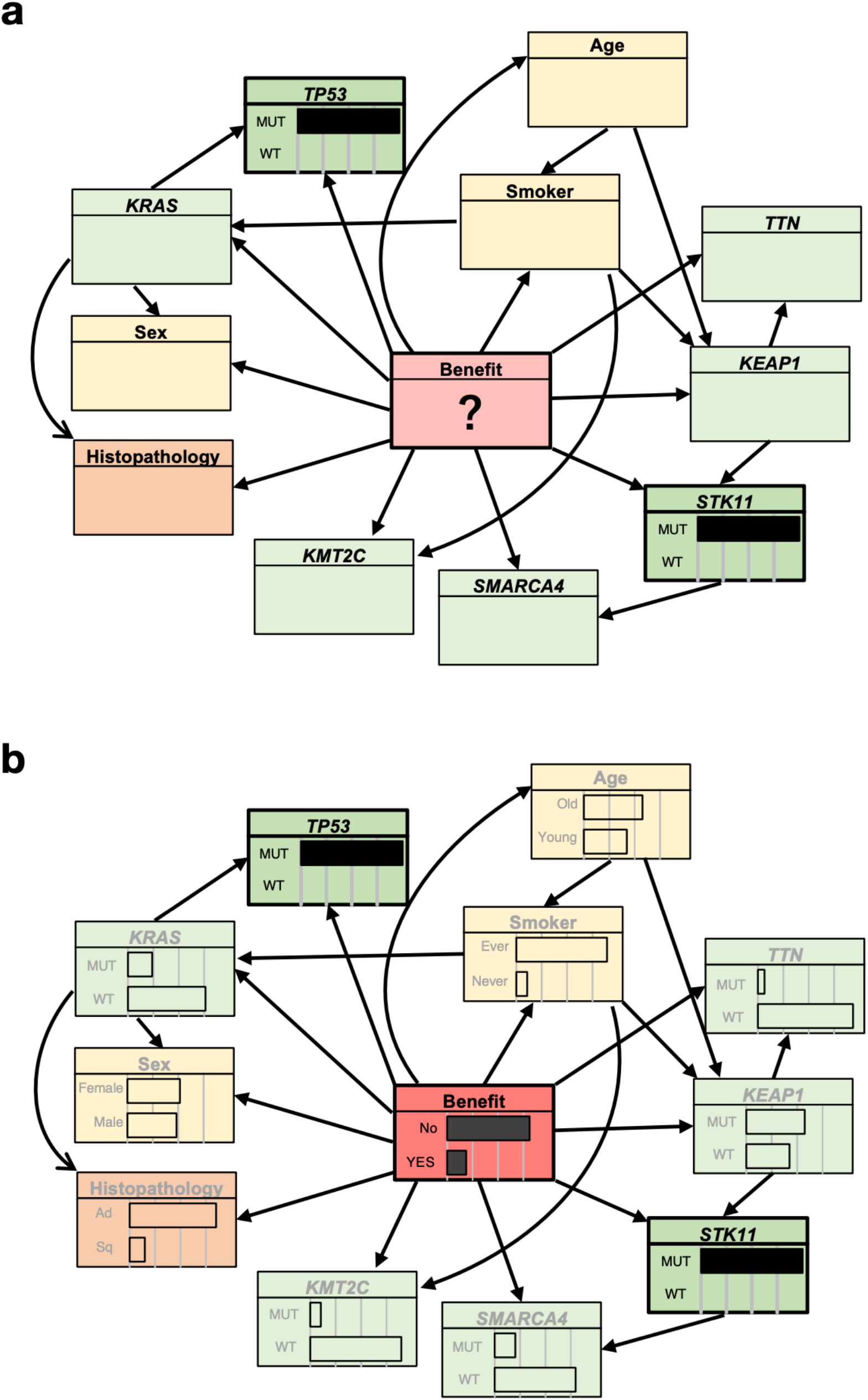
The optimized TAN model can infer DCB even from limited data. (**a, b**) We investigated whether our model (frequency-based TAN, for instance) could infer the durable clinical benefit (DCB) from limited information. (**a**) In this example, the only information provided to the model was the *TP53* and *STK11* mutations in the patient. (**b**) Using rejection sampling, and approximate inference of the probability distribution of the unknown information, we were able to obtain probabilities for all of the hidden states. From only the information that *TP53* and *STK11* are co-mutated, the model computed a response probability to be 0.163, suggesting that immunotherapy would not be effective for this patient, consistent with the previous reports^33^. The white boxes and DCB status were calculated using our approach.

## Discussion

Most prior attempts to predict immunotherapy responses have used ML-based approaches^14– 16^, which are complex “black-box” systems that cannot handle missing data. As input, they require all of the necessary clinical and molecular information to be provided. Such data are often difficult to obtain, especially in hospitals with limited resources, hampering the clinical application of these models.

Transparency and clinical validation are necessary to achieve trustable medical AI^17^. Therefore, we sought to develop an interpretable and robust model that predicts NSCLC patient responses to immunotherapy. We used clinical information, selected mutation data based on the frequency and evidence-based approaches, and established optimized TAN models. Our approach is comparable with, or even superior to, several cutting-edge ML-based methods^15,49,50^, while retaining explainability. It provides clinically informative predictions even when data are limited (Figure 5), as is quite common in clinical settings. Furthermore, because this model only computes conditional probabilities based on Bayes’ theorem^25^, it is possible, if necessary, to control which nodes should have (or should not) have connections, using a “white list” (or “black list”) based on expert knowledge.

We selected several genes based on the mutation frequency or previous evidence. *KRAS*, an immunomodulatory oncogenic gene, leads to escape from immunotherapy^34^. Together with *TP53* or *STK11* mutations, *KRAS* mutation is a potent prognostic factor^33,36^. *STK11* is associated with diminished immunotherapy response^35^. *BRAF* mutation, which are associated with a higher tumor burden, may make tumors vulnerable to immunotherapy^54^. Mutations in driver oncogenes such as *EGFR, ALK*, and *ROS1* in tumors cause a lack of immunogenicity, and thus, a poor response to immunotherapy, regardless of PD-L1 score^55^. Therefore, the expert consensus suggests that patients with these mutations should not be treated with immunotherapy^5^.

Our models could also be used to generate intriguing hypotheses for future research. For instance, our inferences based on limited data (Figure 5) are consistent with the findings of recent reports^33,35^. This suggests that, by using more clinical samples with diverse genetic profiles, our approach may reveal new targets in immunotherapy, providing an invaluable resource for both clinical and basic medicine.

Consistent with an earlier analysis of clinical data on the utility of TAN^48^, our TAN-based approach provided greater value than the NB model. Because of the small sample, the conventional NB model using hill-climbing methods were unable to construct suitable structures for inference (Supplementary Figure 2), suggesting that our approach is better suited to inference with a small dataset. TAN alleviates the conditional constraints imposed by NB. Here, some of the essential connections in TAN structural learning were also detected via model-averaging using bootstrap sampling and MCMC (Figure 3). Our model-averaging findings obtained using the frequency-based approach (Figure 3a, 3b), for instance, strongly suggest an association between smoking status and *KRAS* mutation, which has been established in a previous report^37^. Other strong connections between nodes inferred by model-averaging method (Figure 3) are expected to reveal immunotherapy-related hidden relationships.

In terms of potential limitations, we cannot rule out selection bias due to the integrated use of public datasets. Although the datasets comprise patients who underwent immunotherapy, it is plausible that the data do not represent a specific population. In addition, the strength of the internode relationships that we estimated may reflect the small sample size, and an analysis employing a larger dataset may reveal additional relationships. We therefore developed a web-based intuitive DCB estimator (https://pred-nsclc-ici-bayesian.shinyapps.io/Bayesian-NSCLC/) that does not require computational expertise. Future analyses with larger clinical samples are likely to overcome these limitations, and provide further support for the validity of this approach.

In summary, these robust models are comparable with, or even superior to, other predictive models for immunotherapy. They can predict meaningful and interpretable connections and inferences, even with a limited number of observations. We hope that this model will guide clinicians in selecting NSCLC patients who require immunotherapy, and expect it to be easily applied to other types of cancer.

## Data Availability

All data produced in the present work are contained in the manuscript.

## Abbreviations

AUC: Area under the curve
BN: Bayesian network
DCB: Durable clinical benefit
HC: Hill-climbing method
ICIs: Immune checkpoint inhibitor
ML: Machine learning
MCMC: Markov Chain Monte Carlo
NB: Naïve Bayes
NN: Neural network
NSCLC: Non-small cell lung cancer
TAN: Tree-Augmented naïve Bayes
TMB: Tumor mutational burden

## Acknowledgements

This work was supported by KAKENHI grants from the Japan Society for the Promotion of Science (JSPS) to H.S. (21K17856). We thank our laboratory members for discussion.

## Contributions

H.H. and H.S. designed the project. H.H. contributed to formal analyses and interpretation. H.H. wrote the draft version of the manuscript. H.S. supervised the study and edited the manuscript. All authors contributed to the article and approved the final version.

