## Supplemental Figures 1 and 2. for "Bayesian network enables interpretable and state-of-the-art prediction of immunotherapy responses in cancer patients"

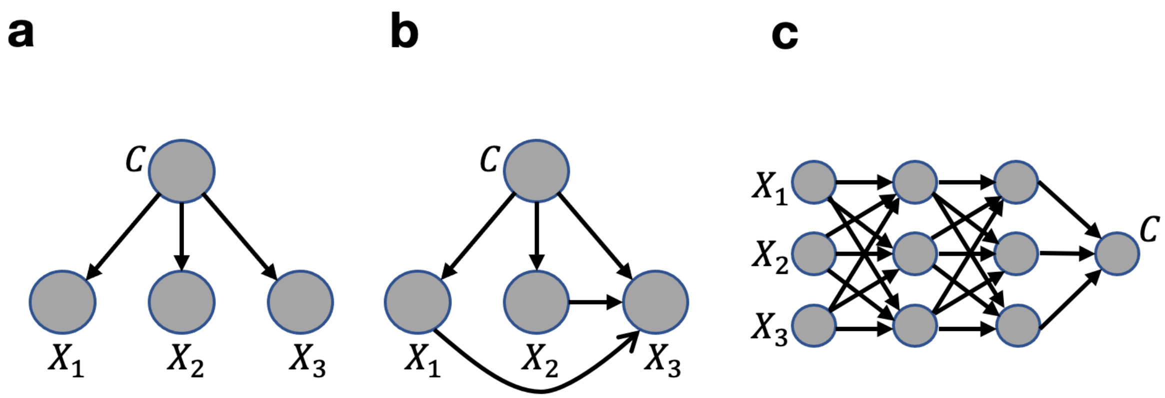


**Supplementary Figure 1. Structures of the naïve Bayes (NB) model, Tree-Augmented naïve Bayes (TAN) model, and a neural network**

(**a**) Naïve Bayes model (NB): the node (an objective variable, indicated by C) is directly connected with covariates (indicated by $X_1, X_2, X_3$). (**b**) Tree-Augmented naïve Bayes (TAN) : unlike the NB model, an additional node can be connected between nodes. (**c**) Neural Network : every node has connections with all other nodes in the previous layers. This illustrates that the NB and TAN models differ substantially from the neural network in their constraints, based on their architecture.

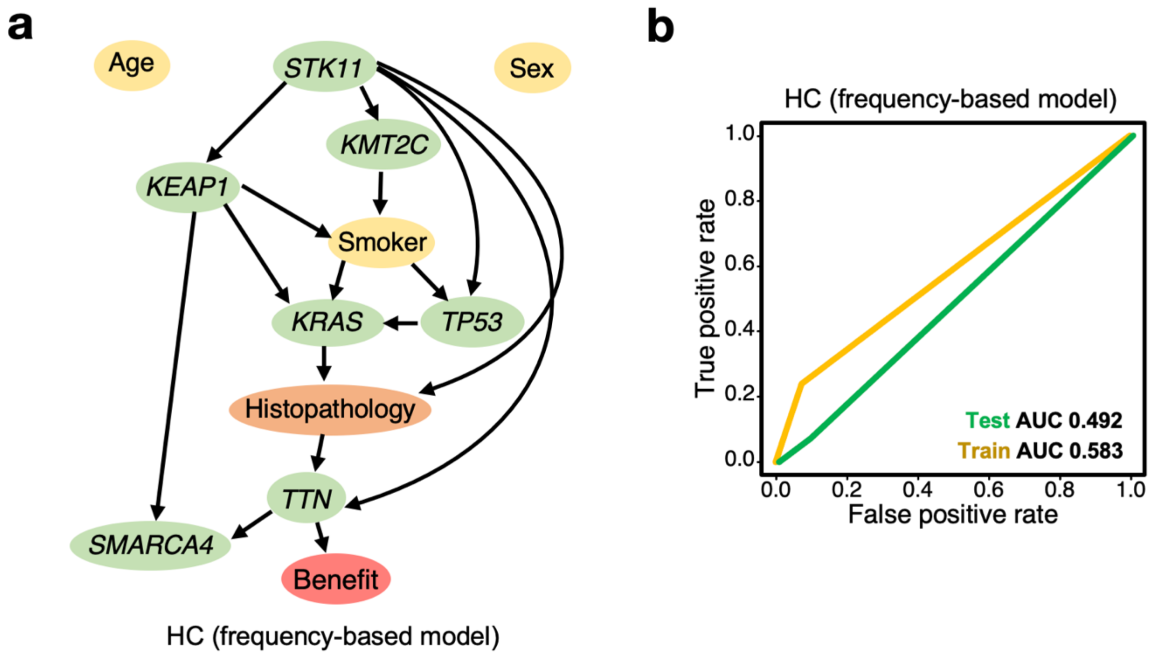


**Supplementary Figure 2. A constraint-free Bayesian network did not achieve better performance**

(**a**) Network structure estimated using an unconstrained Bayesian network model and the hill-climbing method. Note that not all of the variables are connected with edges. (**b**) The unconstrained Bayesian network model in panel (**a**) performed worse, in terms of AUC, than the naïve Bayes (NB) and Tree-Augmented naïve Bayes (TAN) models. For the NB model AUC values, see Figures 2c and 2g; for those of the TAN model, see Figures 2d and 2h.
